# Preventive behavior of Vietnamese people in response to the COVID-19 pandemic

**DOI:** 10.1101/2020.05.14.20102418

**Authors:** Nhan Phuc Thanh Nguyen, Tuyen Dinh Hoang, Vi Thao Tran, Cuc Thi Vu, Joseph Nelson Siewe Fodjo, Robert Colebunders, Michael P. Dunne, Van Thang Vo

**Affiliations:** Institute for Community Health Research, University of Medicine and Pharmacy, Hue University, Vietnam; Faculty of Public Health, University of Medicine and Pharmacy, Hue University, Vietnam; Global Health Institute, University of Antwerp, Belgium; Faculty of Health, Queensland University of Technology, Australia

**Keywords:** COVID-19, Vietnam, social distancing, face masks, adherence

## Abstract

We sought to evaluate the adherence of Vietnamese adults to COVID-19 preventive measures, and gain insight into the effects of the epidemic on the daily lives of Vietnamese people. An online questionnaire survey was organized from March 31 to April 6, 2020. The questionnaire assessed preventive behavior using multiple-answer responses to indicate the extent of adherence. In total, 2175 respondents completed the questionnaire (age range: 18–69 years). The mean adherence scores for personal and community preventive measures were 7.23 ± 1.63 (range 1–9) and 9.57 ± 1.12 (range 1–11), respectively. Perceived adaptation of the community to lockdown (β=2.64, 95% CI 1.25–4.03), fears/worries concerning one’s health (β=2.87, 95% CI 0.04–5.70), residing in large cities (β=19.40, 95% CI 13.78–25.03), access to official COVID-19 information sources (β=16.45, 95% CI 6.82–26.08), and belonging to the healthcare sector (β=22.53, 95% CI 16.00–29.07) were associated with a higher adherence score to anti-COVID instructions. The study indicates excellent preventive behavior of the Vietnamese population which explains the low number of COVID-19 infections and zero recorded mortality up to the first week of May 2020. Further monitoring is recommended to assess the sustainability of COVID-19 prevention via behavior change in the medium and long-term.

## Introduction

In December 2019, an unknown viral pneumonia outbreak erupted in China and has been spreading on a global scale (1). Due to the high likelihood of virus transmission via large droplets and fomites, COVID-19 infection spread rapidly around the planet (2). Preventive public health measures have been implemented to fight the pandemic. Although the strategies applied internationally are similar, the timeliness, scale, and assertiveness of implementation regimes have varied considerably (3).

In Vietnam, the first person with a COVID-19 infection was detected on January 23^rd^ and as of May 5^th^, 2020, Vietnam had totaled 271 confirmed cases with zero deaths (4). Currently Vietnam is among the countries with the lowest number of reported cases, which is remarkable given its population size (approx. 95 million people) and proximity to the epicenter. From the start of the outbreak the government of Vietnam implemented intensive control in the northern Vinh Phuc province (considered to be the local focus of the disease) using a strategy of rapid testing for early detection of sources of infection, assertive contact tracing, timely isolation and free clinical care for people with the infection. Community preventive efforts were implemented early and have been pervasive throughout the country. The government supported social distancing, self-isolation of vulnerable people, mandatory isolation of symptomatic people and those who test positive, focal environmental sanitization, frequent hand washing and wearing of face masks in all public spaces.

By February 25^th^, one month after the first case was recorded, all patients had successfully recovered and had been discharged from hospitals. After more than 20 days with no new case reported, the 17^th^ positive case of COVID-19 was confirmed on March 6^th^. Another wave of the epidemic hit the country with cases being imported from Europe, the USA, and other countries. This led to an increase in domestic transmission of COVID-19, thus ushering in the second stage of the epidemic. Fortunately, the government and health agencies had pandemic preparedness and control plans in place following the fairly recent experience with fighting SARS, Swine Flu and Avian Influenza. The Government implemented national measures restricting travel and suspended visas for foreigners entering Vietnam. On March 20^th^, community transmission was indicated when the 86^th^ and 87^th^ COVID-19 patients had no travel history and no apparent contact with COVID-19 patients (4). To further prevent disease spread in the community, on March 31^st^ the Prime Minister mandated urgent measures, including strict social distancing throughout the country for 15 days. Accordingly, all people were required to stay at home, only go out in case of necessity, and keep a minimum distance of at least 2 meters when moving outdoors; shut down all non-essential business activities and services, only allow essential services such as food distribution, non-elective medical procedures, pharmacies store and the fuel supply. In addition, gatherings of more than 2 people were prohibited (5).

The primary purpose of this study was to assess how well the people have adhered to these instructions because they are crucial in preventing the spread of the virus. We also sought to investigate the effects of the epidemic on the daily lives of Vietnamese people.

## Materials and methods

### Study design

We conducted a descriptive cross-sectional survey during which we received voluntary responses continuously for seven consecutive days (from March 31^st^ to April 6^th^, 2020).

### Study procedures

Data were collected through an online survey initiated by the ICPcovid consortium (https://www.icpcovid.com/). A secure website was used to design and host an online questionnaire, which was adapted to local Vietnamese context. The research team translated the English questionnaire into Vietnamese, conducted a pilot test and improved the questionnaire before official use. It took about 10 minutes to complete the questionnaire. The web link to the online questionnaire was disseminated via various social media platforms, and consenting volunteers submitted their information anonymously. The data became available immediately upon submission. The online questionnaire was kept open for one week (recruitment period) after which it was closed and inaccessible.

### Sample size and sampling

Sampling was done using the snow-ball approach: as more persons completed the online questionnaire, they were encouraged to share the survey web link to their contacts. Only data from respondents who self-identified as being at least 18 years old, who were Vietnamese citizens, understood the Vietnamese language, and resided in Vietnam at the time of the study were retained for this analysis.

### Data collection

The first part of the questionnaire gathered socio-demographic information, including participants’ age, gender, profession, urban vs rural residence, religion, educational level, marital status, housing conditions and household composition.

Given that our study sought to evaluate the level of adherence to the preventive measures recommended by the government, our study outcomes included the proportions of participants who reported following the lockdown instructions against COVID-19 Yes or No answers were given to show whether the person had followed each guideline during the previous week. Overall adherence was assessed by summing the answers with higher scores reflected higher adherence. We also asked participants to self-report their difficulty in staying home as required during the lockdown, using a 5-point Likert scale (1=not difficult at all to 5 =extremely difficult).

Adherence to **personal preventive measures** was assessed by using 9 questions, covering the following aspects: following the 1.5–2m meters social distance rule; wearing a face mask when outside; avoiding touching the face; covering of mouth and nose when coughing/sneezing; hand hygiene via regular hand washing and/or disinfection with sanitizer; frequency of body temperature check; disinfecting mobile phone frequently.

Adherence to **community preventive measures** was assessed with 11 questions with a focus on the following strategies: avoiding meetings/gatherings; avoiding being in a vehicle/bus with more than 10 persons; avoiding going to crowded entertainment venues/ public gym/ beauty salon; avoiding funeral attendance; avoiding going to a fresh food market; usage of individual spoons and plates when eating together with family/non-family members; avoiding travel to another province/country during the lockdown period.

### Study covariates

In addition to socio-demographic characteristics, data were collected to adjust the observed adherence scores for potential confounders. These items inquired about domestic life and professional activities during the COVID-19 pandemic. Fears about the participants’ health as well as their family well-being were measured on a 5-point Likert scale (1= not worried/afraid to 5= extremely worried/afraid). Similarly, perceived adaptation of the community to lockdown instructed from the government was evaluated via 10-point Likert scale (1= no adaption to 10= very strong adaptation).

### Ethical Considerations

Anonymity and informed consent were assured. The study was approved by the Ethical Review Committee of Hue University of Medicine and Pharmacy, Vietnam (No. H202/041 dated March 30^th^, 2020).

### Data analysis

Statistical analysis was performed using SPSS version 20.0. Descriptive statistics presented continuous data as mean ± standard deviation (SD), while categorical variables were presented as percentages. Adherence to anti-COVID-19 measures was the dependent variable of this study. Multiple linear regression model was used to analyze which independent variables associated with squared-transformed adherence scores. 95% confidence intervals and a p-value of less than 0.05 were used for significance testing.

## Results

### Respondent characteristics and their daily activities

A total of 2192 persons completed the online questionnaire. After data cleaning and application of inclusion criteria, 2175 responses were kept. The participants resided in 55/63 provinces of Vietnam. 1054 (48.5%) lived in major municipalities (Ha No, Ho Chi Minh City, Hai Phong, Can Tho, and Da Nang) and 1431 (65.8%) lived in smaller urban or rural areas. The mean age was 31.39 years (SD: 10.66, range: 18–69), and the majority of participants (66.9%) were women. The characteristics of our study participants are summarized in Table 1.

**Table 1.**
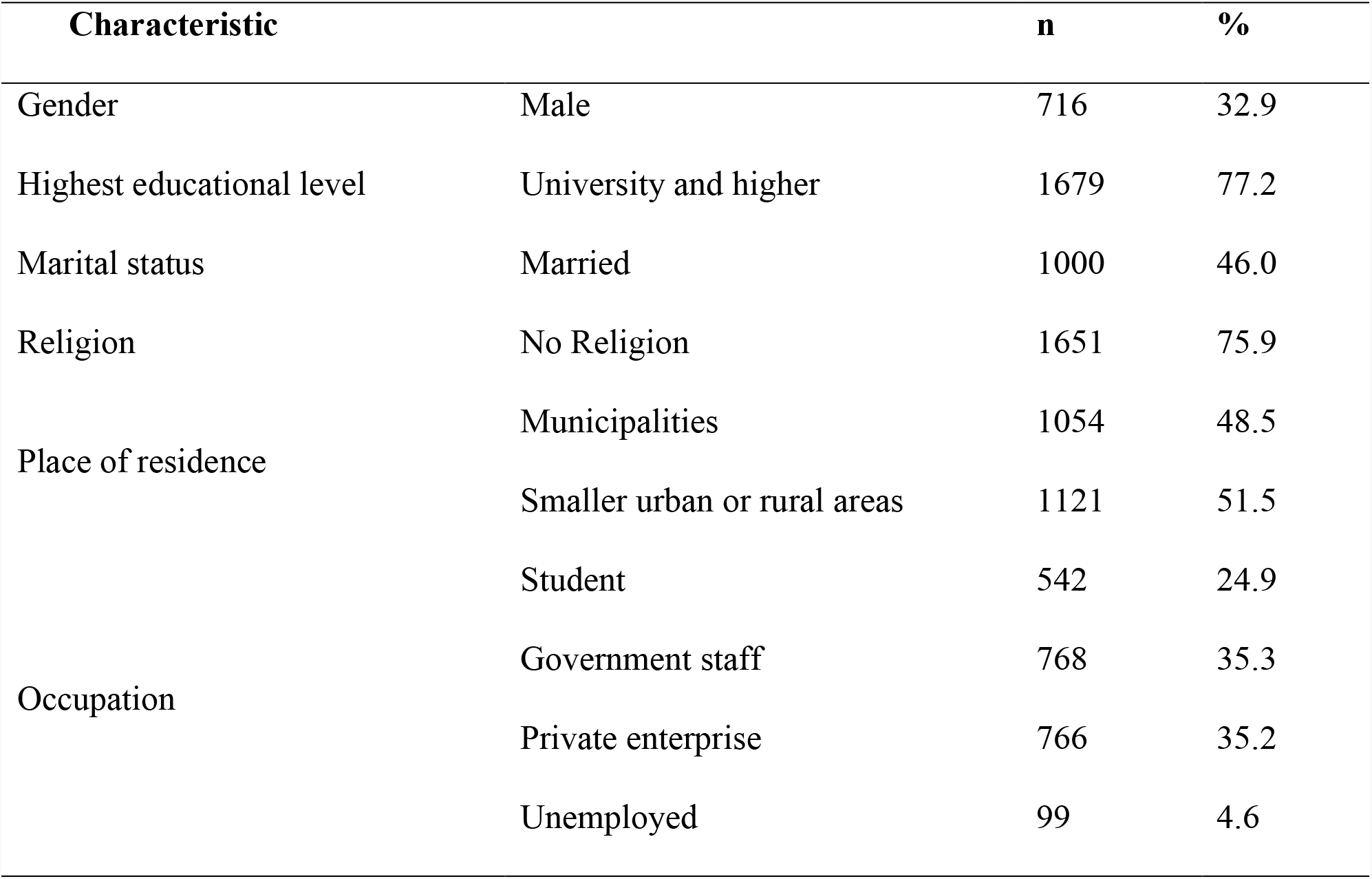
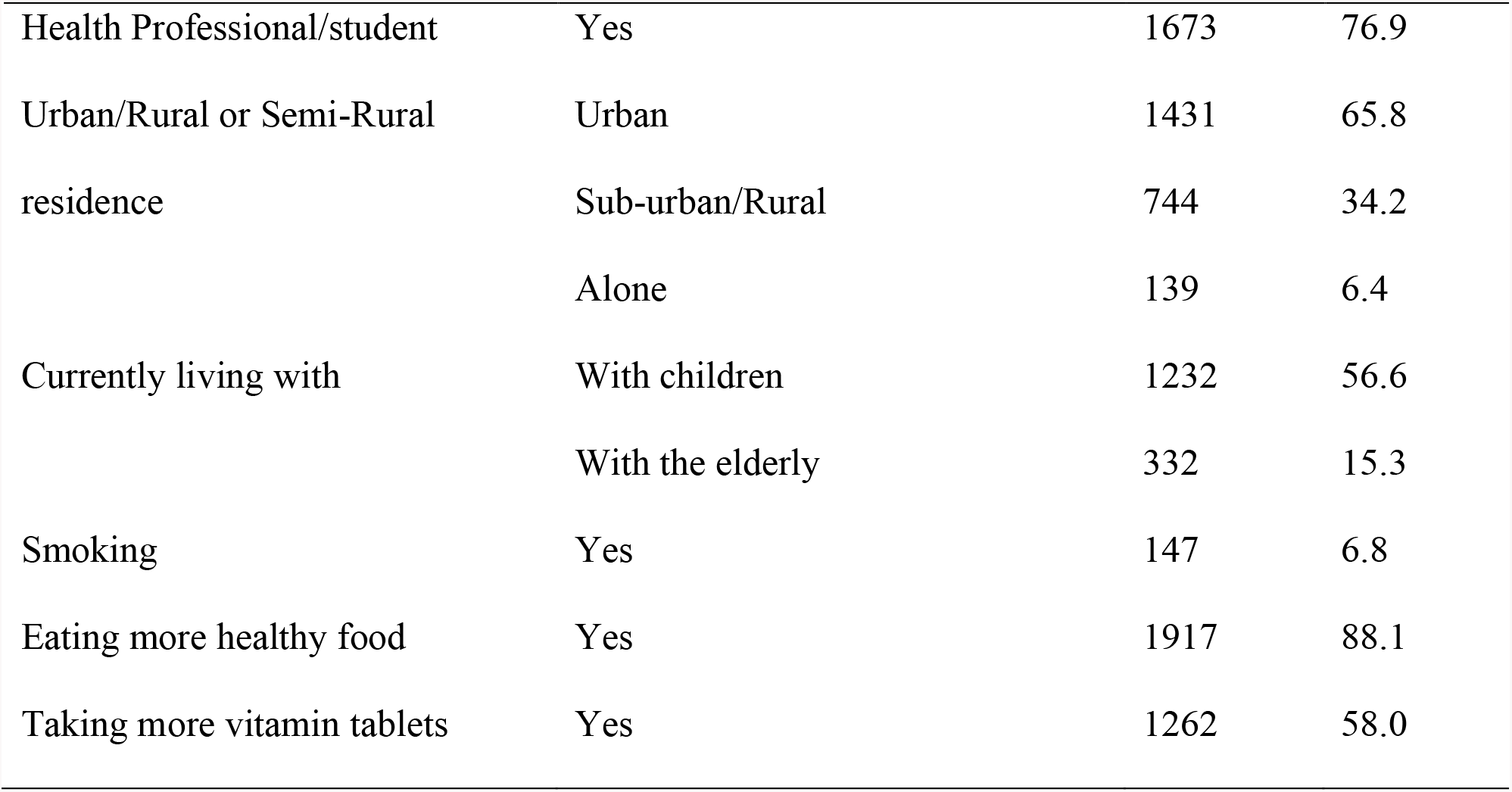
Characteristics of study participants (n=2175) Characteristic.

## Impact of COVID-19 on respondents’ domestic and professional habits

Most participants said they obtained COVID-19 information through official sources such as state television (81.1%) and the Ministry of Health of Vietnam website (74.5%). Of the 1613 participants with a stable job, 777 (48.2%) started working from home because of the epidemic. Confinement measures resulted in 133 (6.1%) participants experiencing difficulties in obtaining food, and 42 (1.9%) persons reported suffering from some form of violence/discrimination because of the restrictive measures taken against COVID-19. Moreover, on a 5-point Likert scale, 30.0% and 42.3% of respondents reported a score of ≥3 indicating that they had moderate to high levels of fear and worry about their own health, and that of their relatives, respectively (Table 2).

**Table 2.**
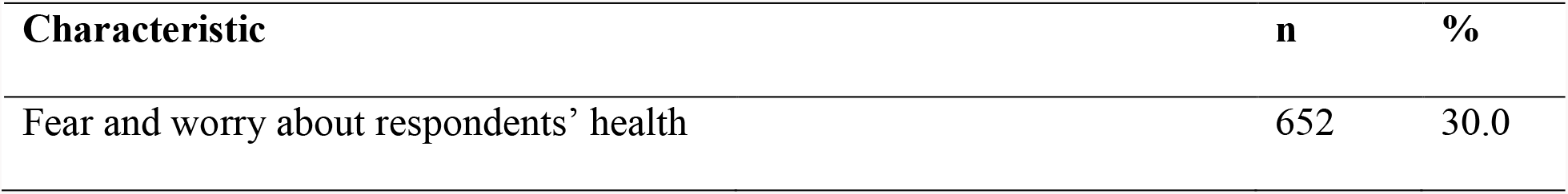
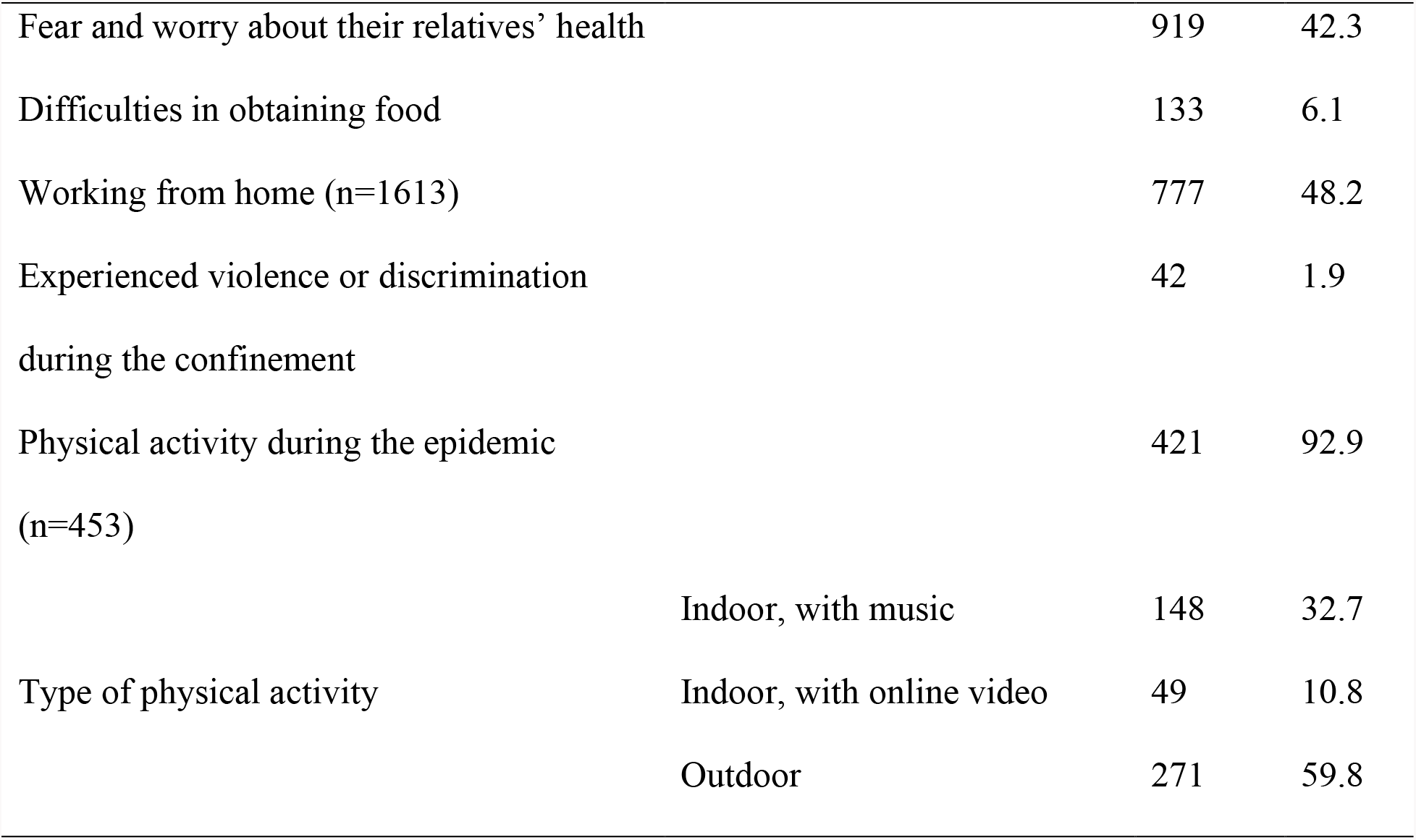
Impact of COVID-19 confinement measures on domestic and professional habits.

## Adherence to social distancing measures in the national response to the threat of COVID-19

### Adherence to personal preventive measures

At the individual level, participants reported few difficulties in complying with the stay-at-home measures instituted by the government (mean difficulty score on the Likert scale: 1.69 ± 0.86; range 1 to 5). “Wearing a face mask when going outside” had the highest compliance rate of 99.5%, “regular hand washing with soap and water during the day” and “covering of mouth and nose with a tissue paper when coughing or sneezing” ranked in the next position, with 97.4% and 94.9%, respectively. The least compliance was found for “measuring body temperature at least twice a week” with 45.1% (Table 3). Using a 9-item score, the mean level of personal adherence to preventive measures was 7.23 ± 1.63; range: 1 to 9.

**Table 3.** Adherence to personal preventive measures for COVID-19.

| Characteristics | n | % |
| --- | --- | --- |
| Follow the 1.5-2m social distance rule | 1919 | 88.2 |
| Face mask use when outdoor | 2165 | 99.5 |
| Cover mouth and nose when coughing/sneezing | 2065 | 94.9 |
| Usually wash/disinfect hands immediately after coughing/sneezing | 1813 | 83.4 |
| Wash hands regularly with water and soap during the day | 2119 | 97.4 |
| Use hand sanitizer/gel regularly during the day | 1767 | 81.2 |
| Body temperature check at least twice a week | 980 | 45.1 |
| Avoid touching my face, eyes, nose and mouth with my hands | 1852 | 85.1 |
| Disinfect phone when I get home | 1047 | 48.1 |

### Adherence to community preventive measures

During the previous week, most of the respondents said they “Had not traveled to another province/country”, “Avoided going to a religious gathering”, and “Avoided going to a public gym” with adherence rates at 99.4%, 99.3% and 99.2%, respectively. However, nearly half of the participants had visited the fresh market in the past seven days (Table 4). Adherence scores for community preventive measures, as assessed by 11 questions, ranged from 0 to 11; mean score: 9.57 ± 1.12. Most respondents reported that their community readily adapted their behaviors to the government’s instructions, as reflected by an adaptability score of ≥6 on the 10-point Likert scale in 76% of cases.

**Table 4.**
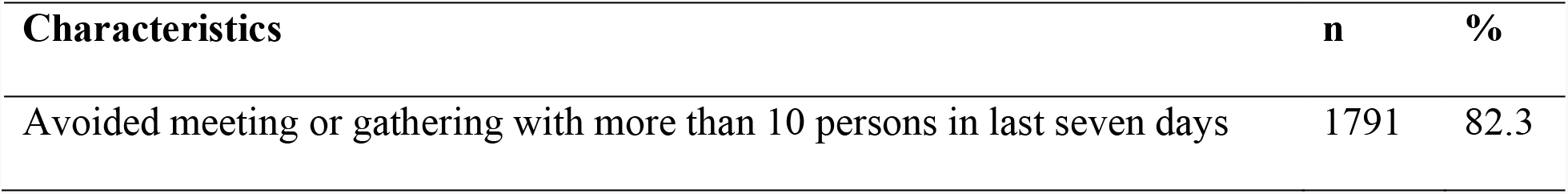
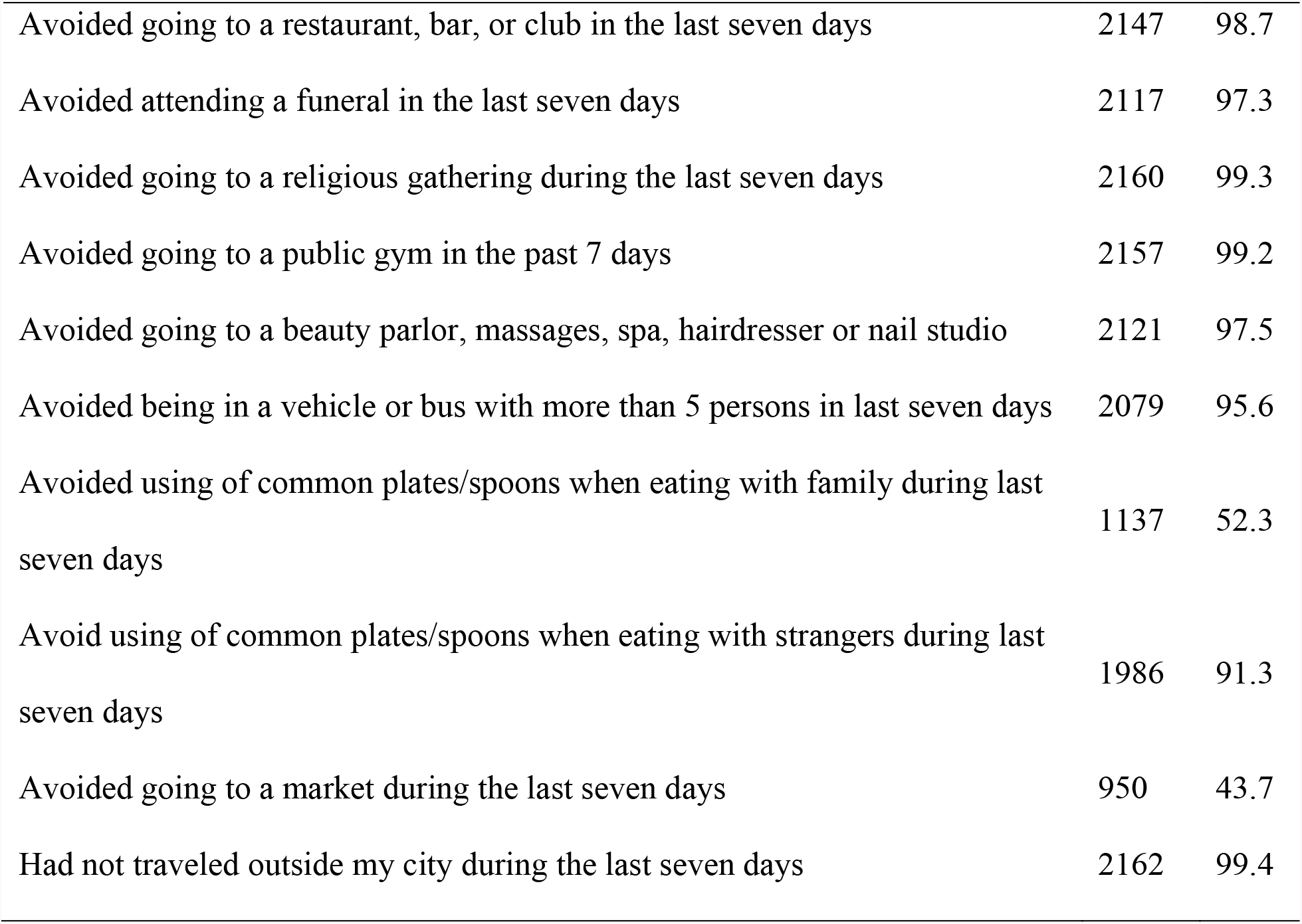
Adherence to community preventive measures for COVID-19.

## Factors associated with adherence to government measures against COVID-19

Summing the responses from self-reports for both personal and community prevention strategies, produced an overall adherence score up to a maximum of 20. Respondents’ scores ranged from 2 to 20, with a mean of 16.80 ± 2.13. Total adherence scores were squared-transformed to fit a normal distribution and used as the dependent variable in linear regression models investigating factors associated with adherence to preventive measures (Table 5). We observed that worries about one’s health, perceived adaptation of the community to the lockdown, residence in large municipalities, official sources of Covid-19 information, and having a professional role in the health sector (worker or student) were associated with higher adherence scores. Conversely, people with higher perceived difficulty in obeying lockdown order to stay at home had significantly lower adherence scores after adjusting for socio-demographic characteristics and other confounders (Model adjusted R-squared = 0.144).

**Table 5.** Multiple linear regression investigating factors associated with adherence to the COVID-19 preventive measures.

| Co-variates | Estimate<br>(95% Confidence interval) | P-value |
| --- | --- | --- |
| Age | -0.08 (-0.36 – 0.20) | 0.567 |
| Gender: Male | 2.86 (-3.09 – 8.81) | 0.346 |
| Fear and worry about their own health (Likert score) | 2.87 (0.04 – 5.70) | 0.047 |
| Perceived adaptation of the community to lockdown (Likert score) | 2.64 (1.25 – 4.03) | <0.001 |
| Difficulty in obeying lockdown (Likert score) | -23.97 (-27.39 – -20.55) | <0.001 |
| Residence in large Municipalities | 19.40 (13.78 – 25.03) | <0.001 |
| Official sources to obtain Covid-19 information | 16.45 (6.82 – 26.08) | 0.001 |
| Being a healthcare worker/student | 22.53 (16.00 – 29.07) | <0.001 |

## Discussion

The government of Vietnam took relatively prompt and intensive measures to reduce the spread of COVID-19 infection in Vietnam. Our data show that most Vietnamese people who participated in the survey complied with most strategies to prevent infection. Very few people resisted the orders for using face masks, frequent hand washing, avoiding large gatherings, or proper social distancing. Probably as a consequence of this early intervention and high uptake of protective behaviors, up to the first week of May 2020 the spread has been minimised with only 63 new cases since the implementation of strategies for the whole population, and no new cases detected in the community since April 16^th^, 2020 (6).

Most companies/organizations have applied unprecedented working methods in accordance with national efforts to promote working from home. This study found that 48.2% of workers were obliged to work from home during the COVID-19 confinement. Although negative effects of social distancing on people’s jobs and lives might emerge if sustained for long periods, the participants in this study indicated relatively few difficulties in the short term, such as meeting daily needs for food.

## Respondents’ adherence to social distancing measures

The survey revealed that many people were moderately to severely worried or afraid about the health risks for family members (42.3%) and this was more than the level of concern about their own health (30.0%). This may reflect the mean age of participants; as most were young adults, they may be concerned about risks to older family members, which is particularly relevant in Vietnam where many people live in multi-generational extended family households.

It is common and easy to apply measures such as wearing a mask, washing hands frequently with soap or disinfectant solutions. Although the efficacy of non-medical masks in preventing COVID-19 spread is currently subject to debate, mask use among infected persons can limit the spread of the virus to the outside environment (7–9). The rate of wearing masks when going out in this study was 99.5%, similar to an estimate of 98% in a Chinese study but higher than 70.1% observed in Japan (9, 10)

Two reasons for such high mask use are the fact that the Vietnamese government made mask use mandatory from April 1^st^, and that in many parts of the country, a majority of the people have a habit of wearing masks to cope with air pollution (5, 11). Although negative social interactions regarding face mask usage have been reported in some parts of the World (12), in Vietnam and some East Asian countries such as China, Japan, and Korea, wearing face masks is ubiquitous (13). It has been practiced for health and cultural reasons (7, 13), so the transition to more widespread mask wearing in response to COVID-19 appears not to have caused a conflict that can sometimes arise if people are forced to change cultural norms.

The implementation of community prevention measures was applied very early in response to a localized outbreak in a northern province, and this was re-enforced from April 1^st^ with official implementation of nationwide social distancing. The shutdown nationally has been unprecedented, with all except essential businesses closed (5). People were advised to stay at home as a patriotic act, and only go out when necessary. Information about outbreaks in healthcare, religious gatherings and entertainment facilities was disseminated widely via mainstream and social media (14, 15). In this survey, the item “Avoided going to a fresh market” had the lowest adherence (43.7%), probably because fresh foods are indispensable in the household and also due to the fact there were many women (66.9%) in the sample and women tend most often to procure fresh food in Vietnam. It is worth noting that in the national social distancing regulations, going to the market is a valid reason to leave the house, although people were asked to reduce the frequency of this activity to the bare minimum (5).

People living in Municipalities had higher adherence scores, perhaps because about 70% of the COVID-19 cases were diagnosed in cities (16). Many respondents were working as healthcare professionals or were medical students, so they may tend to be more adherent to health protection efforts. Age and gender were not significantly associated with adherence score in this study (Table 5).

The high adherence to state recommendations has proven extremely important in the fight against COVID-19 infection. Positive attitudes and compliant behavior indicate that most people in the survey tend to believe in the Government’s motives and requirements, showing solidarity and supportive attitudes during the epidemic. According to Berlin-based Dalia Research, 62% of respondents in Vietnam believe the government is doing the “right amount” in response to the COVID-19 pandemic (17); it is therefore not surprising that Vietnam has been internationally recognized for their success in controlling COVID-19 (18). In our study, the proportion of respondents receiving information from reliable sources was high, which showed that most people were careful to avoid unreliable advice and deliberate misinformation. Notably, the Vietnamese government has sanctioned acts that spread fake news (19).

There are several limitations of this study. First, the respondents were not a representative sample of the Vietnamese population. Indeed, respondents were mainly people from medium to high social strata, since poor and vulnerable populations in Vietnam may have limited internet access. The snowball sampling method and recruitment over just one week possibly explains the high proportion of health professionals and health science students. Random sampling of the population is necessary. Second, it is not possible to verify the veracity of responses provided via a web-based questionnaire. Third, the cross-sectional study design provided only a snapshot of preventive behaviour over one week. It will be important to monitor adherence to official recommendations over time as societies adapt to changing conditions throughout the unpredictable course of this pandemic.

## Conclusion

The study provides insight into compliance with the social distancing and other risk mitigation measures implemented in Vietnam in the context of the COVID-19 pandemic. Overall, adherence to government instructions was high and most likely played a role in rapidly controlling the epidemic in Vietnam and limiting its public health impact. On April 27^th^ the strict lockdown measures were stopped and life is gradually returning to normal in Vietnam, albeit with a stronger than usual emphasis on personal protection during social interactions. Careful monitoring for potential new imported COVID infections and community transmission will be needed to prevent a resurgence of the epidemic.

## Data Availability

All data in the manuscript is available at request

## Acknowledgements

We are grateful to the respondents for their participation. The authors would also like to thank all institutions and stakeholders across the country for supporting us to collect data via online questionnaires. Finally, we would also like to acknowledge Prof. Nguyen Vu Quoc Huy, Rector of University of Medicine and Pharmacy, Hue University, Vietnam for his wonderful support for conducting this study in the difficult time of COVID-19 pandemic occurred.

## Authors’ contributions

TVV, NPTN, JNSF, and RC contributed to the study design and conceptualization. NPTN, TDH and TVV did the statistical analysis, interpretation, data and drafting of the initial manuscript. NPTN and TVV coordinated the study design and data collection. NPTN, TVV, MPD, VTT, CTV, TDH, JNSF and RC critically revised the draft manuscript. All authors have read and approved the final manuscript

